# Enrichment of colibactin-associated mutational signatures in unexplained colorectal polyposis patients

**DOI:** 10.1101/2023.06.02.23290324

**Authors:** D. Terlouw, A. Boot, Q. R. Ducarmon, S. Nooij, M. Suerink, M.E. van Leerdam, D. van Egmond, C.M. Tops, R. D. Zwittink, D. Ruano, A.M.J. Langers, M. Nielsen, T. van Wezel, H. Morreau

## Abstract

Colibactin, a genotoxin produced by polyketide synthase harboring (*pks*^+^) bacteria, induces double-strand breaks and chromosome aberrations. Consequently, enrichment of *pks*^+^ *Escherichia coli* in colorectal cancer and polyposis suggests a possible carcinogenic effect in the large intestine. Additionally, specific colibactin-associated mutational signatures; SBS88 and ID18 in the Catalogue of Somatic Mutations in Cancer database, are detected in colorectal carcinomas. Previous research showed that a recurrent *APC* splice variant perfectly fits SBS88. In this study, we explore the presence of colibactin-associated signatures and fecal *pks* in an unexplained polyposis cohort. Somatic targeted Next-Generation Sequencing (NGS) was performed for 379 patients. Additionally, for a subset of 29 patients, metagenomics was performed on feces and mutational signature analyses using Whole-Genome Sequencing (WGS) on Formalin-Fixed Paraffin Embedded (FFPE) colorectal tissue blocks. NGS showed somatic *APC* variants fitting SBS88 or ID18 in at least one colorectal adenoma or carcinoma in 29% of patients. Fecal metagenomic analyses revealed enriched presence of *pks* genes in patients with somatic variants fitting colibactin-associated signatures compared to patients without variants fitting colibactin-associated signatures. Also, mutational signature analyses showed enrichment of SBS88 and ID18 in patients with variants fitting these signatures in NGS compared to patients without. These findings further support colibactins ability to mutagenize colorectal mucosa and contribute to the development of colorectal adenomas and carcinomas explaining a relevant part of patients with unexplained polyposis.

## Introduction

An enrichment of polyketide synthase (*pks*) encoding *Escherichia coli* in patients with colorectal cancer^1, 2^ and polyposis^3^ has implied a potential carcinogenic effect in the large intestine. These *E. coli* bacteria harbor the *pks* gene island which encodes the necessary equipment to produce the genotoxin colibactin.^4^ Colibactin induces double-strand breaks and chromosome aberrations leading to a specific mutational signature that has been observed in colorectal adenocarcinomas and oral squamous cell carcinomas.^5, 6^ This mutational signature is characterized by T>N mutations with an adenine 3 base pairs to the 5’ side and single thymine deletions located in T homopolymers with 2 to 4 adenines to the 5’ side depending on the length of the T homopolymer. These signatures are documented in the Catalogue of Somatic Mutations in Cancer (COSMIC) database as single base substitution signature SBS88 and indel signature ID18.

*E. coli* is not the only bacterium able to harbor the *pks* gene island. Other bacteria mostly belonging to the Enterobacteriaceae family, such as *Klebsiella pneumoniae, Enterobacter aerogenes* and *Citrobacter koseri*, have also been shown to harbor *pks*.^7^ Moreover, *pks* harboring bacteria are found in other organisms like bacteria in the honey bee gut or a marine sponge.^8^

We previously showed that a common *APC* splice variant c.835-8A>G and several other pathogenic *APC* variants perfectly fit the colibactin-associated mutational signatures.^9^ This finding furthermore implies a possible association between colibactin and the development of colorectal neoplasms. Since a large proportion of our unexplained polyposis patient cohort showed a colibactin-associated *APC* variant in multiple adenomas, further research into the presence and impact of colibactin and its mutational signature was warranted. Therefore, for a subset of polyposis patients, metagenomics was performed on feces and Whole Genome Sequencing (WGS) with subsequent mutational signature analyses was conducted on Formalin Fixed Paraffin Embedded (FFPE) colorectal tissue blocks. Results were compared between those with and without colibactin-associated variants.

## Material and methods

### *APC* mosaicism testing

In total, 379 patients with multiple colorectal adenomas or carcinomas were tested for *APC* mosaicism. In short, DNA was isolated from Formalin Fixed Paraffin Embedded (FFPE) tissue blocks of on average 4 colorectal adenomas or carcinomas using the automated Tissue Preparation System (Siemens). Ampliseq Next Generation Sequencing (NGS) libraries (ThermoFisher Scientific) of a custom-made panel containing 20 colorectal cancer and polyposis associated genes were prepared according to manufacturer’s instructions. Sequencing was performed in an Ion GeneStudio S5 Series sequencer (ThermoFisher Scientific). The raw, unaligned sequencing reads were mapped against human reference genome (hg19) using TMAP software and Torrent Variant Caller was used for variant calling. The detected variants were categorized by pathogenicity and were, when needed, visualized using Integrative Genomic Viewer^10^ or interpreted using the Alamut Visual software (Sophia Genetics).

### *APC* variants and colibactin signature

To determine whether the *APC* variants fit into the mutational signatures SBS88 and ID18, all detected T>N and delT *APC* variants and their sequencing context were visualized using IGV. As previously described^5, 6^, T>N variants with the following sequencing context were labelled as fitting SBS88: 5’ A-(N)-(T/A)-T-(T/A/G) 3’. DelT variants were labelled as fitting ID18 whenever a 2 to 4 adenine homopolymer was flanking the 5’ side of a thymine homopolymer with a total of 5-6 base pairs. As illustrated in figure 1, in 269 patients no somatic variant fitting SBS88 or ID18 was found and therefore served as the control group.

**Figure 1.**
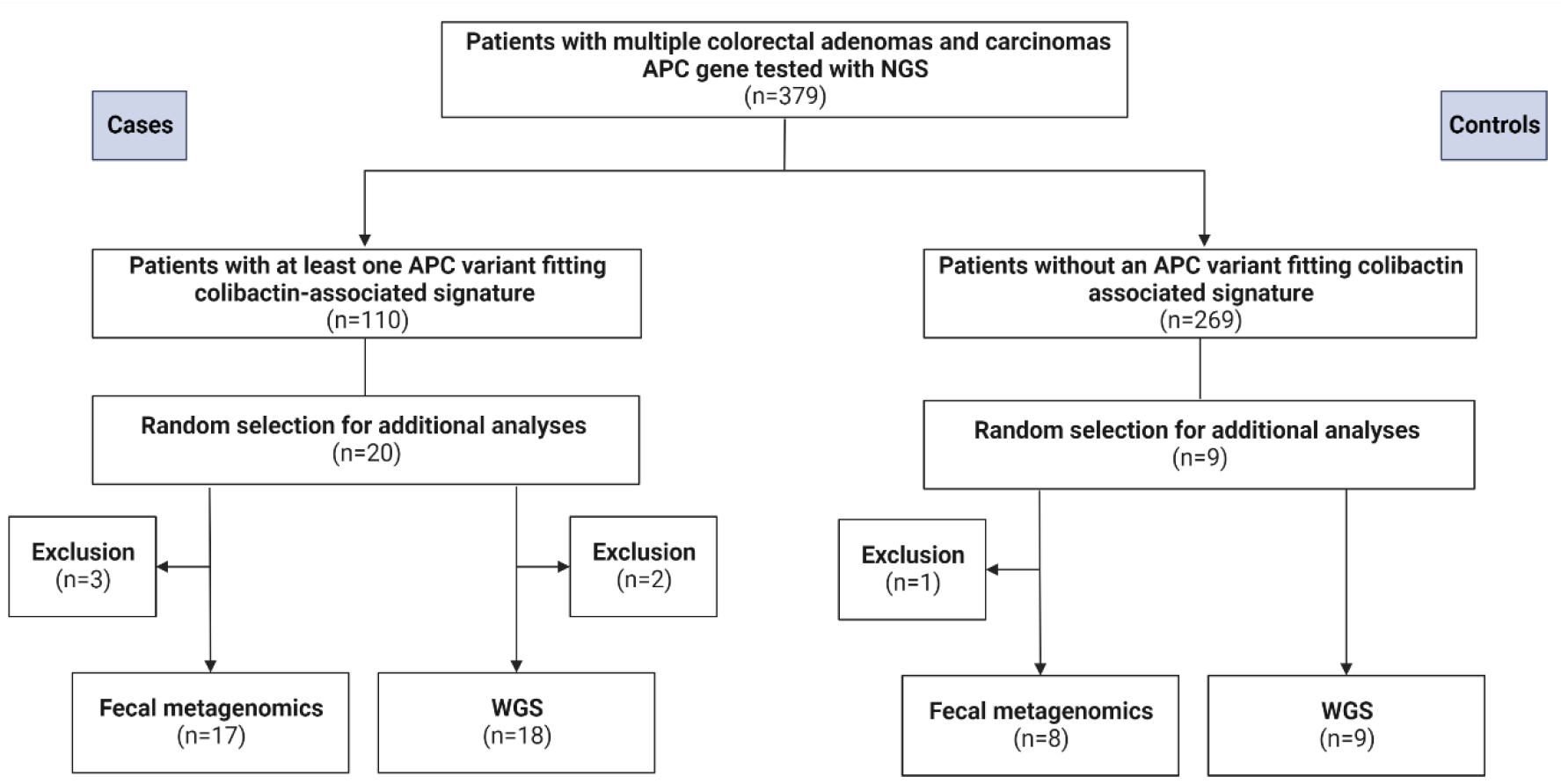
Study design and patient selection. In total, 379 patients were tested using targeted NGS. The case group are patients with at least one *APC* variant fitting colibactin-associated mutational signature. Twenty cases are selected for additional fecal metagenomics and WGS. Patients without APC variant fitting colibactin-associated signatures serve as controls. Nine controls were selected for fecal metagenomics and WGS. Four patients could not be included for fecal metagenomics since they did not respond to sample request (N=3) or passed away (N=1). Two cases were excluded for WGS due to insufficient amount of DNA.

### Case and control group selection

A random selection of twenty-nine patients were included for fecal metagenomics and/or Whole-Genome Sequencing, as depicted in figure 1. Twenty of these patients have adenomas or carcinomas with an *APC* variant suiting SBS88 or ID18 and nine control patients do not have such a colibactin-associated *APC* variant. The patient characteristics are summarized in table 1 and somatic *APC* variants per lesion in supplemental table 1. Furthermore, the sequencing context of the *APC* variants are included in supplemental table 2.

**Table 1.** Phenotypic characteristics and NGS results of patients included for fecal metagenomics and WGS.

| ID | #Ad | Ad age range first | CRC age range first | #SBS88/ID18 | #tested | % | Feces | WGS |
| --- | --- | --- | --- | --- | --- | --- | --- | --- |
| 1 | 22 | 66-70 | 41-45 | 2 | 7 | 28.6 | Y | Y |
| 2 | >9 | 66-70 | 66-70 | 3 | 3 | 100.0 | Y | Y |
| 3 | 15 | 46-50 | 46-50 | 6 | 7 | 85.7 | Y | N |
| 4 | 13 | 66-70 | 66-70 | 3 | 9 | 33.3 | Y | Y |
| 5 | 70 | 66-70 | 66-70 | 3 | 10 | 30.0 | N | Y |
| 6 | 4 | 61-65 | 61-65 | 3 | 6 | 50.0 | Y | Y |
| 7 | 18 | 56-60 | - | 0 | 4 | 0.0 | Y | Y |
| 8 | 10 | 51-55 | 51-55 | 3 | 4 | 75.0 | Y | Y |
| 9 | 2 | 51-55 | 51-55 | 2 | 4 | 50.0 | Y | Y |
| 10 | 10 | 51-55 | - | 8 | 10 | 80.0 | Y | Y |
| 11 | 28 | 81-85 | - | 2 | 6 | 33.3 | Y | N |
| 12 | 36 | 71-75 | - | 2 | 4 | 50.0 | N | Y |
| 13 | 27 | 61-65 | 61-65 | 2 | 3 | 66.7 | Y | Y |
| 14 | 3 | 21-25 | - | 3 | 3 | 100.0 | Y | Y |
| 15 | 10 | 51-55 | - | 2 | 6 | 33.3 | Y | Y |
| 16 | 11 | 51-55 | - | 2 | 3 | 66.7 | Y | Y |
| 17 | 22 | 66-70 | - | 2 | 4 | 50.0 | Y | Y |
| 18 | 14 | 61-65 | - | 2 | 6 | 33.3 | Y | Y |
| 19 | 24 | 41-45 | - | 2 | 4 | 50.0 | N | Y |
| 20 | 10 | 56-60 | - | 2 | 4 | 50.0 | Y | Y |
| 21 | 18 | 46-50 | - | 0 | 4 | 0.0 | Y | Y |
| 22 | 18 | 61-65 | - | 0 | 4 | 0.0 | Y | Y |
| 23 | 20 | 61-65 | - | 0 | 3 | 0.0 | Y | Y |
| 24 | 14 | 46-50 | - | 0 | 4 | 0.0 | Y | Y |
| 25 | 10 | 46-50 | 46-50 | 4 | 4 | 100.0 | Y | Y |
| 26 | 8 | 46-50 | 46-50 | 0 | 4 | 0.0 | Y | Y |
| 27 | 4 | 21-25 | 21-25 | 0 | 4 | 0.0 | N | Y |
| 28 | 2 | 61-65 | 61-65 | 0 | 3 | 0.0 | Y | Y |
| 29 | 9 | 61-65 | 61-65 | 0 | 5 | 0.0 | Y | Y |

### Fecal metagenomics

Feces samples of 25 out of 29 patients were collected for deep fecal shotgun metagenomic sequencing (figure 1 and table 1). Four patients could not be included since they did not respond (N=3) or passed away (N=1). Fecal metagenomic sequencing was performed as previously described.^11^ In short, stool samples were stored at -80°C, DNA was extracted and libraries were prepared according to manufacturer’s protocol.

Sequencing was performed on the Novaseq6000 platform (Illumina, San Diego, CA, USA). Raw metagenomic sequences were processed, analyzed and compared to the *pks* gene island partly comparable to the method description by Nooij et al.^12^ Reads mapping to the human genome (GRCh38) were removed using bowtie2 (version 2.4.2^13^) and SAMtools (version 1.11^14^) and filtered reads were quality-trimmed using fastp (version 0.20.1^15^). The pre-processing workflow is available at (https://git.lumc.nl/snooij/metagenomics-preprocessing). The quality-trimmed reads were screened for the presence of the *pks* island by mapping to the colibactin gene cluster (accession ID AM229678) using BWA-MEM (version 0.7.17^16^). Mapped reads were deduplicated using Picard MarkDuplicates (version 2.23.3^17^) to remove technical artifacts and improve quantification. The *pks* screening workflow is available at (https://git.lumc.nl/snooij/screen_pks_in_polyposis_fecal_metagenomes). As previously outlined, fecal samples positive for at least one *pks* gene were considered *pks*-positive.^12^ To quantify reads per kilobase per million (RPKM) of the individual genes in the *pks* island present in the stool samples, RPKM values were calculated using the following formula: (N of mapped reads/N of base pairs of the coding sequence of the respective gene)*1,000 divided by N of trimmed and filtered reads*1,000,000. For mean RPKM of the entire *pks* island, RPKMs of all individual genes were summed and divided by the total number of 19 *clb* genes.

### Whole-Genome Sequencing (WGS)

DNA from adenomas and carcinomas of 27 out of 29 patients was included for WGS (figure 1 and table 1). Two patients were excluded due to an insufficient amount of DNA extracted.

DNA was isolated from FFPE tissue blocks using the NucleoSpin DNA FFPE XS kit (BIOKE, Leiden, the Netherlands) according to manufacturer’s instructions. WGS was performed on the BGIseq500 platform (BGI, Hong Kong, China) for 4 out of 27 patients (ID 8, 10, 12 and 13). Sequencing for the remaining 23 patients was performed on the NovaSeq6000 platform (Illumina, San Diego, USA). The raw sequencing reads were aligned to a reference genome (GRCh38). The alignment, variant calling and filtering were performed as described before.^6, 18^ The mutational signature assignment using reference mutational signatures was performed using mSigAct::sparseAssignSignatures followed by mSigAct signature presence test, which provides a p-value for the null-hypothesis that a signature is not needed to explain an observed somatic mutation profile compared with the alternative hypothesis that the signature is needed, as previously described.^6^

### BMI and lifestyle data

Body Mass Index (BMI) and information about lifestyle was collected using patient medical records and for some patients using a questionnaire (n=65). BMI was categorized in 4 groups: ≤18.5 ‘underweight’, 18.5-24.9 ‘healthy weight’, 25.0-29.9 ‘overweight’ and ≥30.0 ‘obese’. Both tobacco and alcohol consumption were categorized as ‘never’, ‘former’ and ‘current’. Packyears (PY) was determined as the number of packs of cigarettes smoked per day multiplied by the number of years the patient has smoked.

### Statistical analysis

Statistical analyses were performed using IBM SPSS statistics 25 (Armonk, NY, USA) and a p-value of <0.05 was considered statistically significant. Independent T tests, Chi-square tests and Fisher’s exact tests were used to assess the differences between the patients with and without colibactin variants based on the targeted NGS and patients with *pks* in feces with and without contribution of SBS88 and/or ID18 in the WGS data.

## Results

In total, 379 unexplained polyposis patients were tested for somatic *APC* mosaicism using targeted NGS. In 110 patients, at least one colorectal adenoma or carcinoma harbored an *APC* variant that fits with one of the colibactin-associated mutational signatures. Phenotypic characteristics, like adenoma count, age at first adenoma and personal history of colorectal carcinoma did not significantly differ between the patients with (cases) and without (controls) *APC* variants fitting colibactin mutational signatures. Similarly, lifestyle factors like BMI and smoking status were not significantly different between cases and controls (supplemental table 3). The control group consisted of significantly more former alcohol consumers compared to the cases.

### Fecal metagenomics

Fecal samples from seventeen patients with *APC* variants fitting SBS88 or ID18 (cases) and eight patients without *APC* variants fitting SBS88 or ID18 (controls) were used for metagenomic analysis to detect *pks* genes. As shown in table 2, 59% (10 out of 17) of the cases were *pks* positive compared to 25% (2 out of 8) of controls (p-value=0.124).

**Table 2.** Results of fecal pks using metagenomics and mutational signatures SBS88 and ID18 using WGS.

| ID | % | WGS SBS88 | WGS ID18 | Fecal pks |
| --- | --- | --- | --- | --- |
| 7 | 0 | 0/2 | 0/2 | No |
| 21 | 0 | 0/1 | 0/1 | No |
| 22 | 0 | 0/2 | 0/2 | No |
| 23 | 0 | 1/2 | 0/2 | Yes |
| 24 | 0 | 0/2 | 0/2 | No |
| 26 | 0 | 0/2 | 0/2 | No |
| 27 | 0 | 0/1 | 0/1 |  |
| 28 | 0 | 0/2 | 0/2 | Yes |
| 29 | 0 | 0/2 | 0/2 | No |
| 1 | 28.6 | 1/2 | 0/2 | Yes |
| 5 | 30.0 | 0/2 | 0/2 |  |
| 4 | 33.3 | 0/2 | 0/2 | Yes |
| 11 | 33.3 |  |  | No |
| 15 | 33.3 | 0/2 | 0/2 | Yes |
| 18 | 33.3 | 1/3 | 1/3 | Yes |
| 6 | 50.0 | 2/3 | 0/3 | No |
| 9 | 50.0 | 1/2 | 0/2 | Yes |
| 12 | 50.0 | 0/1 | 0/1 |  |
| 17 | 50.0 | 0/2 | 0/2 | Yes |
| 19 | 50.0 | 0/3 | 0/3 |  |
| 20 | 50.0 | 0/4 | 0/4 | Yes |
| 13 | 66.7 | 0/1 | 0/1 | No |
| 16 | 66.7 | 0/2 | 0/2 | No |
| 8 | 75.0 | 1/1 | 0/1 | Yes |
| 10 | 80.0 | 2/2 | 0/2 | No |
| 3 | 85.7 |  |  | No |
| 2 | 100.0 | 0/3 | 0/3 | Yes |
| 14 | 100.0 | 0/3 | 1/3 | No |
| 25 | 100.0 | 0/3 | 0/3 | Yes |

% - percentage of adenomas or carcinomas tested with NGS with a colibactin variant, WGS SBS88 – number of samples with SBS88 / number of samples tested, WGS ID18 – number of samples with ID18 / number of samples tested, Fecal pks – Yes for patients with and no for patients without pks in their feces sample.

In addition, fecal metagenomics was used to quantify *pks* using RPKM values. However, no significant correlation between number of adenomas/carcinomas with *APC* variants fitting SBS88 or ID18 and the *pks* RPKM values was observed (Pearson: R=0.16, p-value=0.45).

Also, no significant difference in phenotype was observed between the 10 cases with *pks* genes in feces and 7 cases without. When comparing lifestyle factors, a trend was observed towards a higher BMI in the group with *pks* in their feces (supplemental table 4).

None of the bacteria previously associated with colorectal cancer, like *Fusobacterium nucleatum, Bacteroides fragilis, Campylobacter jejuni* and *Clostridioides difficile*, or capable of producing colibactin, like *K. pneumoniae, E. aerogenes* and *C. koseri*, were detected in any of the stool samples (data not shown).

### Mutational signature analysis

For WGS, fifty-seven colorectal adenomas or carcinomas and six normal colon mucosa samples were analyzed from eighteen patients with *APC* variants fitting SBS88 or SBS18 (cases) and nine patients without these variants (controls).

As summarized in table 2, mutational signature analysis identified SBS88 in 8 adenomas or carcinomas derived from 6 cases and ID18 in 2 lesions of 2 cases. Overall, colibactin-associated mutagenesis was detected in 38.9% (7 out of 18) cases. One adenoma of nine controls (11.1%) also showed colibactin associated mutagenesis (SBS88).

### Combining fecal metagenomics and mutational signature analyses

Fifteen cases and eight controls were analyzed both using fecal metagenomics and WGS to compute mutational signature analyses. In 10 cases *pks* was found in their feces samples of which 5 patients also showed a contribution of SBS88 or ID18. Of the 5 cases without *pks* in their feces, three showed SBS88 or ID18 contribution. In 2 controls, *pks* genes were detected in feces and in one of them SBS88 was determined in colorectal lesions. Therefore, 86.7% (13 out of 15) of cases and 25% (2 out of 8) of controls showed hints of *pks* or its carcinogenic effects (p-value=0.006).

No significant differences were detected in lifestyle factors between fecal *pks+* and SBS88/ID18+ cases and fecal *pks+* and SBS88/ID18-cases (supplemental table 5).

## Discussion

Using targeted NGS, 379 patients with unexplained colorectal polyposis were tested for *APC* mosaicism. At least one somatic *APC* variant fitting one of the colibactin associated mutational signatures (SBS88 or ID18) was found in 29% (n=110) patients. Except for the distribution of former alcohol consumption, no significant differences were observed in phenotypic characteristics or lifestyle factors between patients with and without these *APC* variants. Although further research is warranted, the significant difference in former alcohol consumption observed between the groups is likely attributable to the small number of patients with a former alcohol consumption status.

Fecal metagenomics revealed 59% (10 out of 17) of cases with one or more *pks* genes. This proportion is comparable to *pks*^+^ *E. coli* bacteria found in colon mucosa of individuals with Familial Adenomatous Polyposis (68%) and sporadic CRC patients (55%).^1, 3^ In contrast, only 25% (2 out of 8) of controls showed *pks* genes. Although numbers are small, this is comparable to the previously reported incidence of healthy individuals with *pks* genes in feces (27-29%)^12, 19, 20^ and with *pks*+ *E. coli* bacteria in colon mucosa (19-22%).^1, 3^

The current study found no significant differences in phenotypic characteristics and tobacco and alcohol consumption between patients with and without *pks* in feces. Further research is required to draw a conclusion about the correlation between BMI and *pks*^+^ *E. coli*. Although not directly linked to BMI, Arima et al.^21^ found that the association between the Western diet and colorectal cancer patients was only significant in patients with *pks*^+^ *E. coli* in their tumor, suggesting a potential interactive carcinogenic effect between diet and *pks*^+^ *E. coli*.

WGS with subsequent mutational signature analysis showed a contribution of SBS88 or ID18 in 39% (7/18) of cases, compared to 11.1% (1/9) of controls. In only one case all analyzed samples showed a contribution of SBS88. This might be explained by the variable distribution of colonic crypts with the signature within one patient.^22^ Moreover, as summarized in supplemental table 1, the majority of adenomas and carcinomas (n=25) selected for WGS from cases did not harbor *APC* variants fitting SBS88 or ID18. Eighteen of these adenomas and carcinomas were located in the right colon and right sided carcinomas were less likely to have colibactin-associated signatures.^23^

Combining both fecal metagenomics and mutational signature analyses, 86.7% (13/15) of cases showed a significant enrichment towards colibactin influence compared to 25% (2/8) of controls in which both analyses were performed.

This significant enrichment of fecal *pks* and colibactin-associated mutational signatures in cases compared to controls, supports the proposition of a recent preprint that the *APC* splice variant c.835-8A>G might be used as a biomarker for *pks*^+^ *E. coli* influence in the development of the adenoma or carcinoma.^23^

Despite the enrichment, no clear correlation between *pks* in feces and colibactin-associated mutational signatures in colorectal lesions was observed in individual cases. Multiple hypotheses might explain (part of) this finding, comprising both biological and technical issues:

It was previously described that colibactin has a short-term effect, affecting the colon early in life.^22, 24, 25^ Colonic mucosa of patients with a contribution of SBS88 and ID18 might therefore be affected by colibactin, but the *pks*-encoding bacteria may have been eradicated from the intestinal tract at time of feces sampling.

The other way around, in patients with *pks* detected in feces but no SBS88 or ID18 in WGS, enrichment of *pks*^+^ bacteria after the development of adenomas but before feces sampling seems unlikely as *pks*^+^ *E. coli* is detected in feces of newborns and therefore proposed to be transmitted during birth.^25, 26^ These patients might, however, have some kind of mechanism inhibiting colibactin from entering the host cell or whenever inside the cell protects against the specific DNA damage.

The protein ATG16L1 for example is described to be associated with preventing colorectal tumorigenesis in presence of *pks*^+^ *E. coli* in cell lines and mouse models.^27^ Also, colibactin production is in a recent preprint suggested to be inhibited by oxygen.^28^ On the other hand, inflammation seems to promote the expansion of the colibactin-encoding *E. coli* and creates an opportunity to adhere to colon mucosa.^2^ Moreover, co-localization with *B. fragilis* seems to increase DNA damage with faster tumor onset in mice.^3^ These hypotheses might also play a role in whether presence of *pks*+ *E. coli* in the intestinal tract actually leads to DNA damage.

Technically, the small sample set and use of shotgun metagenomics and FFPE tissue blocks are limitations of this study. Especially WGS performed on FFPE samples affects the variant and signature calling and interpretation due to fragmentation and deamination artefacts.^29-31^ Moreover, shotgun fecal metagenomics is a broad analyses but a more sensitive qPCR approach performed at multiple timepoints and at time of adenoma diagnosis could give more insight into the association with adenoma development.

To conclude, in 29% of our cohort with unexplained polyposis patients a colibactin influence was suggested based on targeted NGS data. A subset of cases was included for additional analyses and showed further evidence of colibactin in fecal metagenomics and mutational signature analyses compared to controls. Further research, circumventing the complications of WGS on FFPE tissue and validating the feces analyses, should be performed to draw conclusions for individual cases. Still, these findings provide evidence that colibactin affects the colonic mucosa and plays a pivotal role in unexplained polyposis patients.

## Data Availability

All data produced in the present study are available upon reasonable request to the authors

## Author contributions

DT: acquistion of data, analysis and interpretation of data, drafting the manuscript. AB: performed mutational signature analysis, critical revision of manuscript. QRD, SN: performed fecal metagenomics analysis, critical revision of manuscript. MS: acquisition of data, critical revision of manuscript. MEvL: acquisition of patients, critical revision of manuscript. DvE: performed laboratory work NGS and WGS, critical revision of manuscript. CMT: acquisition of data, critical revision of manuscript. RD: bioinformatics and critical revision of manuscript. RZ: concept fecal metagenomics, critical revision of manuscript. AMJL: acquisition of patients, interpretation of data, critical revision of manuscript. MN: acquisition of patients. TvW, HM, MN: study concept and design, interpretation of data, obtained funding, critical revision of manuscript.

## Ethics

This study was approved by a the ethics review board (B18.042) of the Leiden University Medical Center and all subjects provided written informed consent.

