## Supplemental table for "Enrichment of colibactin-associated mutational signatures in unexplained colorectal polyposis patients"

**Supplemental table 1 -** Somatic variants detected using targeted NGS in the 29 patients selected for additional fecal metagenomics and WGS.

| **ID** | **Type** | **Loc** | **Diff** | **APC #1** | **Cov** | **VAF** | **APC #2** | **Cov** | **VAF** | **APC #3** | **Cov** | **VAF** | **LOH** | **Clb?** | **WGS (Clb)** |
| --- | --- | --- | --- | --- | --- | --- | --- | --- | --- | --- | --- | --- | --- | --- | --- |
| 1 | TA | C | Low | c.2097G>A | >2000 | 0.26 |  |  |  |  |  |  | - |  |  |
|  | TA | C | Low | c.4385delA | 1459 | 0.62 |  |  |  |  |  |  | + |  |  |
|  | TVA | A | Low |  |  |  |  |  |  |  |  |  |  |  | T1 |
|  | TA | D | Low | c.646-1G>A | >2000 | 0.16 | c.2605_2606delAA | >2000 | 0.25 |  |  |  | - |  |  |
|  | TVA | A | Low | c.847C>T | >2000 | 0.32 | c.4260_4261delCA | >2000 | 0.36 |  |  |  | - |  | T2 |
|  | TA | S | Low | c.835-8A>G | >2000 | 0.32 |  |  |  |  |  |  | + |  |  |
|  | TA | S | Low | c.835-8A>G | >2000 | 0.24 |  |  |  |  |  |  | + |  |  |
| 2 | TA | R | Low | c.835-8A>G | 2000 | 0.22 |  |  |  |  |  |  | + |  | T2 |
|  | CRC | S |  | c.835-8A>G | 300 | 0.33 | c.646C>T | >2000 | 0.18 |  |  |  | - |  | T3 |
|  | TA | R | Low | c.835-8A>G | >2000 | 0.28 | c.2417_2418delAT | >2000 | 0.27 |  |  |  | - |  | T1 |
| 3 | TA | A | Low | c.4549C>T | >2000 | 0.18 | c.2008A>T | 483 | 0.29 |  |  |  | - |  |  |
|  | TA | T | Low | c.4549C>T | >2000 | 0.22 | c.2008A>T | 1087 | 0.21 |  |  |  | - |  |  |
|  | TA | S | Low | c.835-8A>G | >2000 | 0.18 | c.2626C>T | >2000 | 0.29 |  |  |  | - |  |  |
|  | TA | R | Low | c.376G>T | 513 | 0.32 | c.1313-10_1313-9delTAinsAG | 1781 | 0.29 |  |  |  | - |  |  |
|  | TA | T | Low | c.835-8A>G | >2000 | 0.17 |  |  |  |  |  |  |  |  |  |
|  | TA | T | Low | c.4063delT | >2000 | 0.11 |  |  |  |  |  |  |  |  |  |
|  | TA | S | Low | c.835-8A>G | >2000 | 0.22 | c.3595_3596delAA | 1941 | 0.24 |  |  |  | - |  |  |
| 4 | CRC | C |  |  |  |  |  |  |  |  |  |  |  |  | T1 |
|  | TA | D | Low | c.835-8A>G | >2000 | 0.16 |  |  |  |  |  |  | - |  |  |
|  | TA | D | Low | c.694C>T | 357 | 0.2 | c.4108A>T | >2000 | 0.19 |  |  |  |  |  |  |
|  | TA | S | Low | c.835-8A>G | >2000 | 0.69 |  |  |  |  |  |  | + |  |  |
|  | TA | T | Low | c.1409-5A>G | 1533 | 0.22 | c.2222delA | 455 | 0.25 |  |  |  | - |  |  |
|  | TA | T | Low | c.1409-5A>G | 640 | 0.26 | c.2222delA | 1993 | 0.23 |  |  |  | - |  |  |
|  | TA | D | Low | c.2977A>T | 1998 | 0.26 | c.646-1G>A | 996 | 0.25 |  |  |  | - |  |  |
|  | TA | D | Low | c.694C>T | 49 | 0.37 |  |  |  |  |  |  |  |  | T2 |
|  | TA | D | Low | c.3951delA | 627 | 0.54 |  |  |  |  |  |  |  |  |  |
| 5 | CRC | S |  | c.835-8A>G | >2000 | 0.21 |  |  |  |  |  |  |  |  |  |
|  | AD | UNK | NS | c.1690C>T | >2000 | 0.22 |  |  |  |  |  |  |  |  |  |
|  | AD | UNK | NS | c.835-8A>G | >2000 | 0.31 | c.637C>T | >2000 | 0.25 |  |  |  |  |  |  |
|  | AD | UNK | NS | c.835-8A>G | >2000 | 0.29 |  |  |  |  |  |  |  |  |  |
|  | AD | UNK | NS | c.1690C>T | >2000 | 0.2 | c.3473_3474delGA | >2000 | 0.23 |  |  |  | - |  |  |
|  | TVA | UNK | Low | c.3581C>G | >2000 | 0.43 | c.637C>T | >2000 | 0.42 |  |  |  | - |  | T1 |
|  | TA | UNK | Low | c.694C>T | 165 | 0.77 |  |  |  |  |  |  | + |  |  |
|  | TA | UNK | Low | c.2802_2805delTTAC | 1919 | 0.29 |  |  |  |  |  |  | - |  |  |
|  | TVA | UNK | Low | c.3709_3710delCA | >2000 | 0.73 |  |  |  |  |  |  | + |  | T2 |
|  | TVA | R | Low | c.3438_3439delTT | >2000 | 0.79 |  |  |  |  |  |  | + |  |  |
| 6 | CRC | R |  | c.4285C>T | >2000 | 0.65 | c.3070C>G | >2000 | 0.6 |  |  |  | + |  |  |
|  | CRC | R |  | c.1483delA | 398 | 0.23 |  |  |  |  |  |  | - |  | T1 |
|  | TVA | D | Low | c.2309C>A | >2000 | 0.39 | c.4466delT | >2000 | 0.34 |  |  |  |  |  | T2 |
|  | TA | D | Low | c.2051T>C | >2000 | 0.58 | c.6868T>C | 738 | 0.12 |  |  |  | + |  |  |
|  | TA | S | Low | c.835-8A>G | >2000 | 0.73 |  |  |  |  |  |  | + |  |  |
|  | TA | S | Low | c.4501delT | >2000 | 0.42 |  |  |  |  |  |  | + |  | T3 |
| 7 | TA | A | Low |  |  |  |  |  |  |  |  |  |  |  | T1 |
|  | TA | A | Low |  |  |  |  |  |  |  |  |  |  |  |  |
|  | TA | HF | Low | c.637C>T | >2000 | 0.16 | c.5673G>C | 165 | 0.18 |  |  |  | - |  |  |
|  | TA | T | Low |  |  |  |  |  |  |  |  |  |  |  | T2 |
| 8 | SSL | T | Low | c.835-8A>G | >2000 | 0.24 |  |  |  |  |  |  |  |  |  |
|  | TVA | D | Low | c.4063delT | 1665 | 0.28 | c.4464_4465delAT | >2000 | 0.5 |  |  |  | - |  |  |
|  | TVA | S | Low | c.1495C>T | 242 | 0.19 |  |  |  |  |  |  | - |  | T1 |
|  | CRC | S |  | c.835-8A>G | 1660 | 0.4 | c.646C>T | 774 | 0.27 |  |  |  | - |  |  |
|  | N | D |  |  |  |  |  |  |  |  |  |  |  |  | N |
| 9 | CRC | A |  | c.4666delA | 646 | 0.29 | NM_000038.6:c.2167G>C | 735 | 0.29 |  |  |  | + |  | T1 |
|  | CRC | S |  | c.835-8A>G | 784 | 0.44 |  |  |  |  |  |  | + |  |  |
|  | CRC | RS |  | c.1495C>T | 530 | 0.8 |  |  |  |  |  |  | + |  | T2 |
|  | CRC | R |  | c.835-8A>G | 614 | 0.39 |  |  |  |  |  |  | + |  |  |
| 10 | TA | A | Low | c.1600A>T | >2000 | 0.27 | c.423-6A>G | 1364 | 0.6 |  |  |  | - |  |  |
|  | TA | T | Low | c.4348C>T | >2000 | 0.41 | c.835-8A>G | >2000 | 0.42 |  |  |  | - |  |  |
|  | TA | D | Low | c.4348C>T | >2000 | 0.14 | c.835-8A>G | >2000 | 0.41 |  |  |  | + |  |  |
|  | TA | S | Low | c.835-8A>G | 1708 | 0.67 |  |  |  |  |  |  | + |  |  |
|  | TA | A | Low | c.4053_4070delinACAA | >2000 | 0.69 |  |  |  |  |  |  | + |  | T1 |
|  | TA | D | Low | c.835-8A>G | >2000 | 0.44 |  |  |  |  |  |  | + |  |  |
|  | TA | R | Low | c.835-8A>G | >2000 | 0.35 | c.646C>T | >2000 | 0.22 |  |  |  | - |  |  |
|  | TA | D | Low | c.835-8A>G | >2000 | 0.35 | c.3404_3405delAT | >2000 | 0.28 |  |  |  | - |  |  |
|  | TA | UNK | Low | c.1600A>T | >2000 | 0.37 | c.423-6A>G | 1622 | 0.58 |  |  |  | - |  |  |
|  | TA | R | Low |  |  |  |  |  |  |  |  |  |  |  | T2 |
|  | N | T |  |  |  |  |  |  |  |  |  |  |  |  | N |
| 11 | TA | C | Low | c.835-8A>G | 958 | 0.1 |  |  |  |  |  |  | - |  |  |
|  | TA | D | Low | c.646C>T | >2000 | 0.26 |  |  |  |  |  |  | - |  |  |
|  | TVA | S | Low |  |  |  |  |  |  |  |  |  |  |  |  |
|  | TA | T | Low | c.835-8A>G | 808 | 0.08 |  |  |  |  |  |  | - |  |  |
|  | TA | A | Low | c.3472A>T | >2000 | 0.23 |  |  |  |  |  |  | - |  |  |
|  | TA | T | Low | c.3907C>T | 291 | 0.4 |  |  |  |  |  |  | - |  |  |
| 12 | TA | HF | Low |  |  |  |  |  |  |  |  |  |  |  | T1 |
|  | TA | T | Low | c.835-8A>G | 63 | 0.14 | c.694C>T | 63 | 0.25 |  |  |  | - |  |  |
|  | TA | D | Low | c.835-8A>G | 157 | 0.29 |  |  |  |  |  |  | + |  |  |
|  | TA | R | Low | c.3867delT | 1772 | 0.57 |  |  |  |  |  |  | + |  |  |
|  | N | HF |  |  |  |  |  |  |  |  |  |  |  |  | N |
| 13 | TVA | D | Low | c.835-8A>G | 160 | 0.35 | c.694C>T | 263 | 0.28 |  |  |  | - |  |  |
|  | CRC | S |  | c.3493A>T | >2000 | 0.3 | c.3577_3578delCA | 194 | 0.26 |  |  |  | - |  |  |
|  | TA | T | Low | c.1660C>T | 439 | 0.1 |  |  |  |  |  |  | - |  | T1 |
|  | N | T |  |  |  |  |  |  |  |  |  |  |  |  | N |
| 14 | TA | A | Low | c.835-8A>G | 862 | 0.49 |  |  |  |  |  |  | + |  | T1 |
|  | TA | S | Low | c.835-8A>G | 438 | 0.74 |  |  |  |  |  |  | + |  | T2 |
|  | TA | S | Low | c.835-8A>G | 548 | 0.27 | c.1549-8A>G | 771 | 0.20 |  |  |  | - |  | T3 |
| 15 | TA | UNK | Low | c.4477_4478dupAC | 1937 | 0.18 | c.4062_4066delTTCTT | 697 | 0.55 |  |  |  | - |  |  |
|  | TA | UNK | Low | c.835-8A>G | >2000 | 0.32 | c.1449_1450delTG | >2000 | 0.26 |  |  |  | - |  |  |
|  | TA | R | Low | c.835-8A>G | 571 | 0.12 | c.637C>T | 1012 | 0.11 |  |  |  | - |  |  |
|  | TVA | UNK | NS | c.4099C>T | 181 | 0.56 |  |  |  |  |  |  | - |  |  |
|  | TA | R | Low | c.1495C>T | 1889 | 0.17 | c.4067_4070delinsAGA | 824 | 0.3 |  |  |  | - |  | T1 |
|  | TA | D | Low | c.4099C>T | 422 | 0.11 | c.4063dupT | 420 | 0.26 |  |  |  | - |  | T2 |
| 16 | TA | S | Low |  |  |  |  |  |  |  |  |  |  |  | T1 |
|  | TA | S | Low | c.4348C>T | >2000 | 0.36 | c.2442delT | 404 | 0.23 |  |  |  | - |  | T2 |
|  | TA | S | High | c.835-8A>G | 601 | 0.25 | c.778C>T | >2000 | 0.19 |  |  |  | - |  |  |
| 17 | TA | A | Low |  |  |  |  |  |  |  |  |  |  |  | T1 |
|  | TA | HF | Low | c.835-8A>G | 1556 | 0.67 |  |  |  |  |  |  | + |  |  |
|  | TA | T | Low | c.835-8A>G | >2000 | 0.43 | c.1342_1349delCCTGCTGT | 603 | 0.18 |  |  |  | - |  |  |
|  | TA | T | Low | c.637C>T | >2000 | 0.31 | c.4391_4394delAGAG | >2000 | 0.33 |  |  |  | - |  | T2 |
|  | TA | HF | Low |  |  |  |  |  |  |  |  |  |  |  |  |
|  | TA | T | Low | c.835-8A>G | 1986 | 0.26 |  |  |  |  |  |  | + |  |  |
|  | TA | T | Low |  |  |  |  |  |  |  |  |  |  |  |  |
| 18 | TA | A | Low | c.3916G>T | 392 | 0.5 |  |  |  |  |  |  |  |  | T1 |
|  | TA | T | Low | c.3927_3931delAAAGA | 224 | 0.54 |  |  |  |  |  |  |  |  |  |
|  | TA | S | Low | c.3916G>T | 655 | 0.65 | c.646C>T | 254 | 0.21 |  |  |  |  |  | T2 |
|  | TA | S | Low | c.835-8A>G | 461 | 0.21 | c.1312+1G>A | 750 | 0.14 |  |  |  |  |  |  |
|  | TA | D | Low |  |  |  |  |  |  |  |  |  | - |  |  |
|  | TA | S | Low | c.646C>T | 423 | 0.15 | c.1627-8A>G | >2000 | 0.27 |  |  |  | - |  | T3 |
| 19 | TA | HF | Low | c.3523C>T | 127 | 0.21 |  |  |  |  |  |  | - |  | T1 |
|  | TA | SF | Low |  |  |  |  |  |  |  |  |  |  |  |  |
|  | TA | D | Low | c.2626C>T | 1089 | 0.23 | c.1312+3A>G | 407 | 0.32 |  |  |  | - |  | T2 |
|  | TA | S | Low | c.4612_4613delGA | 1923 | 0.34 | c.2861T>A | 1997 | 0.36 |  |  |  | - |  | T3 |
| 20 | TA | A | Low | c.4348C>T | 1999 | 0.14 | c.1693G>T | 1996 | 0.18 |  |  |  | - |  | T1 |
|  | TA | T | Low |  |  |  |  |  |  |  |  |  |  |  | T2 |
|  | TA | D | Low | c.835-8A>G | 1985 | 0.67 |  |  |  |  |  |  | + |  | T3 |
|  | TA | S | Low | c.835-8A>G | 1285 | 0.69 |  |  |  |  |  |  | + |  | T4 |
| 21 | TA | A | Low | c.1213C>T | 840 | 0.21 | c.1538delT | 726 | 0.19 | c.4429C>T | 417 | 0.1 | - |  |  |
|  | TA | T | Low | c.1213C>T | 668 | 0.14 | c.734C>A | 196 | 0.31 | c.4348C>T | 1104 | 0.22 | - |  |  |
|  | TA | D | Low | c.646C>T | 178 | 0.19 |  |  |  |  |  |  | - |  |  |
|  | TA | RS | Low | c.1312+5G>T | 1992 | 0.24 | c.637C>T | 1983 | 0.23 |  |  |  | - |  | T1 |
|  | N | D |  |  |  |  |  |  |  |  |  |  |  |  | N |
| 22 | TA | D | Low |  |  |  |  |  |  |  |  |  |  |  | T1 |
|  | TA | D | Low |  |  |  |  |  |  |  |  |  |  |  | T2 |
|  | TA | A | Low |  |  |  |  |  |  |  |  |  |  |  |  |
|  | TA | T | Low | c.4301delG | 1925 | 0.34 | c.1495C>T | 1204 | 0.35 |  |  |  |  |  |  |
| 23 | TVA | C | Low | c.4285C>T | 1229 | 0.25 | c.4090delA | 420 | 0.22 |  |  |  |  |  | T1 |
|  | TVA | D | Low | c.664C>T | 301 | 0.22 | c.2138C>G | 1654 | 0.25 |  |  |  |  |  | T2 |
|  | TA | R | Low |  |  |  |  |  |  |  |  |  |  |  |  |
| 24 | TA | A | Low | c.6019T>C | 988 | 0.52 |  |  |  |  |  |  |  |  | T1 |
|  | TA | T | Low | c.6019T>C | 691 | 0.51 | c.637C>T | 1015 | 0.21 |  |  |  |  |  |  |
|  | TA | S | Low | c.6019T>C | 803 | 0.47 | c.4348C>T | 1465 | 0.25 | c.2821G>T | 1036 | 0.15 |  |  |  |
|  | TA | S | Low | c.6019T>C | 1880 | 0.51 | c.694C>T | 1683 | 0.36 |  |  |  |  |  | T2 |
| 25 | TA | S | Low | c.3088A>T | >2000 | 0.06 | c.4734_4741dup | 1558 | 0.07 |  |  |  | - |  | T1 |
|  | TA | S | Low | c.3088A>T | >2000 | 0.03 | c.3095C>G | 1995 | 0.12 | c.4734_4741dup | 1297 | 0.05 | - |  |  |
|  | TA | S | Low | c.1600A>T | 1999 | 0.24 | c.4314_4315dupAC | 1433 | 0.44 |  |  |  | - |  | T2 |
|  | CRC | R |  | c.4059delA | 680 | 0.16 | c.3880C>T | 286 | 0.17 |  |  |  | - |  | T3 |
| 26 | CRC | T |  | c.4348C>T | 1988 | 0.36 |  |  |  |  |  |  | - |  |  |
|  | TA | A | Low | c.4359delT | 1517 | 0.37 | c.2626C>T | 1412 | 0.39 |  |  |  | - |  | T1 |
|  | TA | S | Low | c.3863delG | 786 | 0.35 | c.694C>T | 1389 | 0.29 |  |  |  | - |  |  |
|  | TVA | S | High |  |  |  |  |  |  |  |  |  | - |  | T2 |
| 27 | AD | S | Low | c.3934G>T | 223 | 0.26 | c.4116_4117dup | 340 | 0.42 |  |  |  | - |  |  |
|  | TA | S | Low |  |  |  |  |  |  |  |  |  | - |  |  |
|  | TA | S | Low | c.3934G>T | 1028 | 0.20 | c.4116_4117dup | 521 | 0.17 |  |  |  | - |  | T1 |
|  | CRC | R |  |  |  |  |  |  |  |  |  |  | - |  |  |
|  | N | R |  |  |  |  |  |  |  |  |  |  |  |  | N |
| 28 | CRC | FH |  |  |  |  |  |  |  |  |  |  |  |  | T1 |
|  | CRC | S |  | c.4099C>T |  |  |  |  |  |  |  |  |  |  | T2 |
|  | AD | S | Low |  |  |  |  |  |  |  |  |  |  |  |  |
| 29 | CRC/TA | D |  | c.3905delT |  |  |  |  |  |  |  |  |  |  |  |
|  | CRC/TA | D |  | c.3905delT |  |  |  |  |  |  |  |  |  |  |  |
|  | CRC/TA | D |  | c.2626C>T |  |  |  |  |  |  |  |  |  |  |  |
|  | CRC/TA | D |  | c.3907C>T |  |  | c.1978_1981dupAACT |  |  |  |  |  |  |  | T1 |
|  | CRC/TA | D |  | c.4309A>T |  |  |  |  |  |  |  |  |  |  | T2 |

Type - type of lesion, TA - tubular adenoma, TVA - tubulovillous adenoma, CRC - carcinoma, AD - adenomatous not further specified, SSL - sessile serrated lesion, N - normal colon mucosa, Loc - location in colon, C - coecum, A - ascendens, HF - hepatic flexure, T - transversum, SF - splenif flexure, D - descendens, S - sigmoid, RS - rectosigmoid, R - rectum, UNK - not specified. Diff - differentiation, Cov - coverage, VAF - Variant Allel Frequency, LOH - Loss of Heterozygosity, Clb? - does 1 of the variants suit SBS88 or ID18, red – no, green – yes. WGS (Clb) – samples selected for WGS, red – no SBS88 or ID18 found, green – SBS88 or ID18 found.

**Supplemental table 2 –** Sequencing context of somatic *APC* variants detected using targeted Next Generation Sequencing.

| ***APC* Variant** | **Seq. context** | **Rev complement** | **Colibactin?** |
| --- | --- | --- | --- |
| c.1312+3A>G | CAAGT**A**TGTTC | GAACA**T**ACTTG | Yes |
| c.1409-5A>G | TTTAA**A**TTAGG | CCTAA**T**TTAAA | No |
| c.1483delA | ACAGT**A**TTACA | TGTAA**T**ACTGT | No |
| c.1538delT | AGATG**T**AGCCA |  | No |
| c.1549-8A>G | ATTTA**A**TTTAC | GTAAA**T**TAAAT | Yes |
| c.1600A>T | AACTA**A**AATCT | AGATT**T**TAGTT | Yes |
| c.1627-8A>G | GATTA**A**TTTGC | GCAAA**T**TAATC | Yes |
| c.2008A>T | ACTTA**A**AATCT | AGATT**T**TAAGT | Yes |
| c.2051T>C | AACTT**T**GTGGA |  | No |
| c.2222delA | ATGCC**A**ATATT | AATAT**T**GGCAT | No |
| c.2442delT | GACAA**T**TTTAA |  | Yes |
| c.2861T>A | CAAAT**T**AGAAT |  | Yes |
| c.2977A>T | AAAGT**A**AGTTT | AAACT**T**ACTTT | Yes |
| c.3088A>T | GTCTT**A**AATAT | ATATT**T**AAGAC | Yes |
| c.3472A>T | AAGAG**A**GACCA | TGGTC**T**CTCTT | No |
| c.3493A>T | GCATA**A**AATAT | ATATT**T**TATGC | Yes |
| c.3867delT | GGATG**T**AATCA |  | No |
| c.3905delT | TACCC**T**GCAAA |  | No |
| c.3951delA | AGCTG**A**AGATC | GATCT**T**CAGCT | No |
| c.4059delA | TGTTG**A**ATTTT | AAAAT**T**CAACA | Yes |
| c.4063delT | TTGAA**T**TTTCT |  | Yes |
| c.4090delA | CCTCC**A**AAAGT | ACTTT**T**GGAGG | No |
| c.4108A>T | CACCC**A**AAAGT | ACTTT**T**GGGTG | No |
| c.423-6A>G | AAAAA**A**AATAG | CTATT**T**TTTTT | Yes |
| c.4309A>T | GAAGT**A**AAACA | TGTTT**T**ACTTC | No |
| c.4359delT | GTACC**T**AAAAA |  | No |
| c.4385delA | TGCTG**A**AAAGA | TCTTT**T**CAGCA | No |
| c.4466delT | CTTTA**T**TACAT |  | No |
| c.4501delT | ATGGA**T**TTTCT |  | Yes |
| c.4666delA | GGCAG**A**AAAAAC | GTTTTT**T**CTGCC | No |
| c.6868T>C | AGACA**T**CCCAA |  | Yes |
| c.835-8A>G | GCTTA**A**TTTTT | AAAAA**T**TAAGC | Yes |

Colibactin? – Yes, whenever a single nucleotide variant matches SBS88 or a deletion aligns ID18.

**Supplemental table 3** – Phenotypic characteristics comparison between patients with at least one colibactin *APC* variant and patients without a colibactin *APC* variant detected in targeted NGS.

|  | **Total** | **No colibactin variant** | **At least 1 colibactin variant** | ***p-value*** |
| --- | --- | --- | --- | --- |
| **Total** | 379 | 269 | 110 |  |
| **Male – n (%)** | 254 | 178 (66.2%) | 76 (69.1) | 0.632 |
| **Adenoma count – n (%)** |  |  |  | 0.446 |
| **0** | 6 | 6 (2.2) | 0 (0.0) |  |
| **1-9** | 46 | 33 (12.3) | 13 (11.8) |  |
| **10-19** | 158 | 108 (40.1) | 50 (45.5) |  |
| **20-29** | 93 | 63 (23.4) | 30 (27.3) |  |
| **30-49** | 52 | 40 (14.9) | 12 (10.9) |  |
| **50-99** | 15 | 11 (4.1) | 4 (3.6) |  |
| **>100** | 9 | 8 (3.0) | 1 (0.9) |  |
| **Age at adenoma diagnosis – mean (min-max)** | 58.6 (17-84) | 58.6 (17-84) | 58.5 (23-83) | 0.931 |
| **CRC, yes – n (%)** | 107 | 70 (26.0) | 37 | 0.166 |
| **Age at (first) CRC diagnosis – mean (min-max)** | 58.3 (25-84) | 57.6 (25-84) | 59.8 (39-75) | 0.361 |
| **BMI – n (%)** | 135 | 92 | 43 | 0.905 |
| Underweight | 2 | 1 (1.1) | 1 (2.3) |  |
| Healthy | 47 | 31 (33.7) | 16 (37.2) |  |
| Overweight | 61 | 43 (46.7) | 18 (41.9) |  |
| Obese | 25 | 17 (18.5) | 8 (18.6) |  |
| **BMI – mean (min-max)** | 26.8 (16.5-48.4) | 26.9 | 26.6 | 0.713 |
| **Smoking – n (%)** | 229 | 153 | 76 | 0.869 |
| Never | 68 | 46 (30.1) | 22 (28.9) |  |
| Former | 82 | 53 (34.6) | 29 (38.2) |  |
| Current | 79 | 54 (35.3) | 25 (32.9) |  |
| **Smoking PY – mean (min-max)** | 14.5 (0-73) | 13.4 | 16.4 | 0.357 |
| **Alcohol – n (%)** | 219 | 147 | 72 | 0.031 |
| Never | 38 | 27 (18.4) | 11 (15.3) |  |
| Former | 12 | 12 (8.2) | 0 (0.0) |  |
| Current | 169 | 108 (73.5) | 61 (84.7) |  |
| **Glasses of alcohol/week – mean (min-max)** | 15.0 (0-168) | 14.5 | 16.1 | 0.668 |

**Supplemental table 4** – Phenotypic characteristics comparison between patients with and without pks genes in their feces using fecal metagenomic sequencing.

|  | **Total** | ***Pks* in feces** | **No *pks* in feces** | ***p-value*** |
| --- | --- | --- | --- | --- |
| **Total** | 17 | 10 | 7 |  |
| **Male – n (%)** | 10 | 7 (70.0) | 3 (42.9) | 0.350 |
| **Adenoma count – n (%)** | 13.6 (2-28) | 13.2 (2-22) | 14.1 (4-28) | 0.813 |
| **Age at adenoma diagnosis – mean (min-max)** | 58.6 (24-81) | 60.7 (49-69) | 55.6 (24-81) | 0.414 |
| **CRC, yes – n (%)** | 8 | 6 (60.0) | 2 (28.6) | 0.335 |
| **Age at (first) CRC diagnosis – mean (min-max)** | 57.9 (42-69) | 56.0 (42-69) | 63.5 (62-65) | 0.362 |
| **BMI – n (%)** | 25.9 (18-33.3) | 27.9 (21.5-33.3) | 22.3 (18-28.1) | 0.041 |
| **Smoking PY – mean (min-max)** | 24.2 (0-73) | 25.2 (0-51) | 22.7 (0-73) | 0.818 |
| **Glasses of alcohol/week – mean (min-max)** | 21.6 (0-140) | 29.5 (6-140) | 11.2 (0-35) | 0.355 |

**Supplemental table 5** – Phenotypic characteristics comparison between patients with pks genes in their feces and SBS88 and/or ID18 in WGS and patients with pks genes in their feces but no SBS88 or ID18 in WGS.

|  | **Total** | **SBS88/ID18+** | **SBS88/ID18-** | ***p-value*** |
| --- | --- | --- | --- | --- |
| **Total** | 10 | 4 | 6 |  |
| **Male – n (%)** | 10 | 2 (50.0) | 5 (83.3) | 0.500 |
| **Adenoma count – n (%)** | 13.6 (2-28) | 12.0 (2-22) | 14.0 (10-22) | 0.650 |
| **Age at adenoma diagnosis – mean (min-max)** | 58.6 (24-81) | 60.5 (55-69) | 60.8 (49-69) | 0.948 |
| **CRC, yes – n (%)** | 6 | 3 (75.0) | 3 (50.0) | 0.571 |
| **Age at (first) CRC diagnosis – mean (min-max)** | 57.9 (42-69) | 50.7 (42-55) | 61.3 (49-69) | 0.232 |
| **BMI – n (%)** | 25.9 (18-33.3) | 27.6 (21.5-33.3) | 28.1 (23.7-32.7) | 0.898 |
| **Smoking PY – mean (min-max)** | 25.2 (0-51 | 19.8 (4-33) | 28.9 (0-51) | 0.425 |
| **Glasses of alcohol/week – mean (min-max)** | 21.6 (0-140) | 16.7 (11-24) | 37.2 (6-140) | 0.573 |
